# Predicting Prognosis and IDH Mutation Status for Patients with Lower-Grade Gliomas Using Whole Slide Images

**DOI:** 10.1101/2021.01.21.21250241

**Authors:** Shuai Jiang, George J. Zanazzi, Saeed Hassanpour

## Abstract

We developed end-to-end deep learning models using whole slide images of adults diagnosed with diffusely infiltrating, World Health Organization (WHO) grade 2 gliomas to predict prognosis and the mutation status of a somatic biomarker, isocitrate dehydrogenase (IDH) 1/2. The models, which utilize ResNet-18 as a backbone, were developed and validated on 296 patients from The Cancer Genome Atlas (TCGA) database. To account for the small sample size, repeated random train/test splits were performed for hyperparameter tuning, and the out-of-sample predictions were pooled for evaluation. Our models achieved a concordance- (C-) index of 0.715 (95% CI: 0.569, 0.830) for predicting prognosis and an area under the curve (AUC) of 0.667 (0.532, 0.784) for predicting IDH mutations. When combined with additional clinical information, the performance metrics increased to 0.784 (95% CI: 0.655, 0.880) and 0.739 (95% CI: 0.613, 0.856), respectively. When evaluated on the grade 3 gliomas TCGA dataset, which was not used for training, our models were able to predict survival with a C-index of 0.654 (95% CI: 0.537, 0.768) and IDH mutations with an AUC of 0.814 (95% CI: 0.721, 0.897). If validated in a prospective study, our method could potentially assist clinicians in managing and treating patients with diffusely infiltrating gliomas.

## INTRODUCTION

Diffuse gliomas are the most common primary brain tumors in adults. According to the World Health Organization (WHO) classification of tumors of the central nervous system^1^, the diffusely infiltrating gliomas are categorized into grade 1 to 4 based on histologic features such as mitotic activity, tumor cell pleomorphism and the presence of necrosis and/or microvascular proliferation^1^. Diffusely infiltrating low grade gliomas (LGGs) refer to grade 2 gliomas, which includes astrocytomas and oligodendrogliomas. High grade gliomas (HGGs) include grade 3 and grade 4 gliomas (glioblastomas, GBM). Over time, all LGGs progress to higher grade. A pair of slightly different terms, low**er**-grade gliomas and high**er**-grade gliomas, are also often used to refer grade 2/3 gliomas, and GBM, respectively. GBM is the most aggressive type of diffuse gliomas with a 5-year overall survival rate of less than 5%^2^ while the lower-grade gliomas usually have better prognosis with the median survival time of more than 7 years^3^.

Isocitrate dehydrogenase (IDH) mutations are common in lower-grade gliomas with a prevalence of about 80%^4^. IDH mutations also occur frequently in secondary GBMs (progressed from lower-grade gliomas) but less so in primary GBMs^5^. IDH mutations are associated with better prognosis among patients with GBMs^5^ and lower-grade gliomas^3,6^.

Treatment methods for diffuse gliomas include surgical resection at the time of diagnosis and subsequent radiation and/or chemotherapy. Recent retrospective studies and clinical studies suggest the presence of an IDH mutation is an important treatment indicator. For example, previous studies showed that extensive surgical resection is beneficial for patients with IDH-mutant astrocytomas^7–10^. Some studies found radiotherapy is more effective than temozolomide monotherapy for the treatment of patients with IDH-mutant 1p/19q non-codeleted tumors^11,12^, and the effectiveness of photon beam radiation therapy in treating patients with IDH-mutant gliomas is being investigated by an ongoing clinical trial^13^. As for patients with IDH wild-type tumors, it has been proposed that conventional radiation therapy might work better given the more aggressive spread of gliomas lacking IDH mutations^14^. All of these underscore the importance of IDH mutation status in clinical treatment planning.

Predicting survival times for adult diffusely infiltrating gliomas is of great interest in clinical practice, which can inform treatment and shared-decision making between physicians and patients. Prognostic factors for adult diffuse gliomas include age, gender, performance status, extent of tumor resection, and intrinsic factors of the tumor including grade, IDH mutation, chromosome 1p/19q status, and MGMT promoter methylation^15,16^. Although tumor tissues are graded according to well-established histological criteria, this process is time-intensive and it can be challenging to utilize this information for survival estimation. The development of deep learning models over the past few years provides unique opportunities to extract information from unstructured data such as whole slide images^17–19^. Several studies have used whole slide images to predict prognosis of patients diagnosed with diffuse gliomas and have shown promising results. Mobadersany et al.^11^ predicted survival of patients diagnosed with grade 2 to 4 gliomas from the TCGA database, and they obtained a C-index of 0.741 in the testing phase. Another study done by Rathore et al.^20^ using all grade 2 to grade 4 gliomas from the TCGA dataset achieved an even higher C-index (0.79 to 0.85 for different subtypes). However, it is not clear what type of loss function was used and how loss to follow-up was treated in this study. A recent study focused on grade 4 gliomas. The authors split the survival times into four categories and applied the cross-entropy loss function. A C-index which is commonly used to measure survival model performance, was not reported in that study^21^.

Inferring IDH mutation status from histological images has also attracted interests over recent years. Despite the importance of IDH mutations in the clinical context, interrogating IDH mutational status can be time-consuming and expensive. However, if the IDH mutation information can be obtained from frozen section slides following surgery, both time and cost can be greatly reduced. A few studies have investigated whether whole slide images can be used for IDH mutation prediction. Momeni et al. applied deep recurrent attention models using TCGA dataset and obtained an AUC of 0.86^22^. With a dataset combining 200 TCGA grade 2 to 4 cases and 66 private cases, an AUC of 0.920 was achieved in predicting IDH mutations^23^.

Although there have been studies on the application of deep learning models using whole slide images of brain tumor patients, information on the performance of deep learning models among patients with lower grades is limited. A model trained using data across grade 2 to grade 4 cases can perform well in distinguishing high-risk patients from low-risk patients with different grade of gliomas; however, it might not be able to differentiate high risk patients from low risk patients within the same grade. In this study, we set our focus on grade 2 gliomas (LGGs) and explored the use of deep learning models for survival prediction and IDH mutation status prediction utilizing the TCGA dataset. This is a more challenging task in comparison to previous studies due to the smaller sample size and more uniform cases. To mitigate this challenge, in our study, we use an ensemble deep learning framework similar to the established method proposed by Wulczyn et al.^24^. We also train and validate the models in repeated random splits of the dataset to obtain pooled out-of-sample predictions, a similar method adopted by Mobadersany et al.^11^, to ensure the stability and quality of our results. This approach helps to obtain the distribution of the model performance in the whole dataset and make the results not subject to unbalanced splitting.

## RESULTS

### Model performance and comparison with clinical features for prognosis prediction

The average performance of the models with the chosen hyperparameters was 0.644 (standard deviation = 0.107) in the 32 separate test splits. The ensembled predictions achieved a C-index of 0.715 (95% CI: 56.9, 83.0) for the prognosis prediction task over the entire dataset (Table 1). Several demographic and clinical variables were considered for survival analysis in our study. A Cox-proportional hazards model of age achieved a C-index of 0.745. When our WSI risk scores were added to the Cox model, the C-index was improved to 0.765 (difference = 0.020, 95% CI: −0.091, 0.100). Gender and race were unrelated to survival with C-index close to 0.5. The C-index when using only a clinical variable (primary diagnosis) was 0.572. By adding our WSI-based risk scores, the C-index was increased to 0.689 (difference 0.117, 95% CI: −0.003, 0.230) but still lower than WSI risk scores alone. IDH mutation status was another strong predictor with C-index of 0.692 without WSI risk scores or 0.762 with WSI risk scores. When combining age and IDH mutations, the C-index was 0.774 (95% CI: 0.658, 0.863), and adding our WSI-based risk scores improved the C-index slightly to 0.784 (difference = 0.010, 95% CI: −0.097, 0.085).

**Table 1.** Model performance statistics for survival prediction task and IDH mutation prediction task, evaluated among patients with LGG. 95% confidence intervals were derived from 10,000 bootstrapping replications. **Bold** texts indicate best performance for each column. * indicates statistically significant difference (p < 0.05).

| Survival prediction performance: C-index [95% CI] |  |  |  |
| --- | --- | --- | --- |
|  | Without WSI Risk Score | With WSI Risk Score | Difference |
| None | - | 0.715 [0.569, 0.830] | - |
| Age | 0.745 [0.627, 0.838] | 0.765 [0.643, 0.865] | 0.020 [-0.091, 0.100] |
| Gender | 0.509 [0.345, 0.630] | 0.688 [0.528, 0.815] | 0.179 [-0.007, 0.352] |
| Race | 0.520 [0.444, 0.554] | 0.713 [0.568, 0.831] | 0.193 [0.056, 0.322]* |
| Primary diagnosis | 0.572 [0.437, 0.707] | 0.689 [0.539, 0.822] | 0.117 [0.003, 0.230]* |
| IDH mutations | 0.692 [0.573, 0.807] | 0.762 [0.602, 0.878] | 0.070 [-0.048, 0.161] |
| Age+IDH mutations | **0.774 [0.658, 0.863]** | **0.784 [0.655, 0.880]** | 0.010 [-0.097, 0.085] |
| IDH mutation prediction performance: AUC [95% CI] |  |  |  |
|  | Without WSI Predicted IDH Mutation Probability | With WSI Predicted IDH Mutation Probability | Difference |
| None | - | 0.667 [0.532, 0.784] | - |
| Age | 0.689 [0.552, 0.816] | 0.726 [0.599, 0.845] | 0.037 [-0.053, 0.097] |
| Gender | 0.536 [0.430, 0.643] | 0.650 [0.507, 0.769] | 0.114 [-0.078, 0.240] |
| Race | 0.567 [0.480, 0.650] | 0.687 [0.560, 0.800] | 0.120 [-0.002, 0.229] |
| Primary diagnosis | 0.519 [0.389, 0.641] | 0.637 [0.472, 0.755] | 0.118 [-0.090, 0.261] |
| Age + Race | **0.711 [0.585, 0.834]** | **0.739 [0.613, 0.856]** | 0.028 [-0.051, 0.078] |

Figure 1a and Figure 1c shows both age and WSI risk score can successfully identify high risk patients (age > 46 years or WSI risk score > 1.37) shortly after diagnosis. However, patients with intermediate risk were not significantly different from patients with low risk. Survival curves for patients with IDH mutations apparently separated with the survival curve for patients with IDH wild-type about half-year post diagnosis (Figure 1b). Log-rank tests were significant for all three predictors.

**Figure 1.**
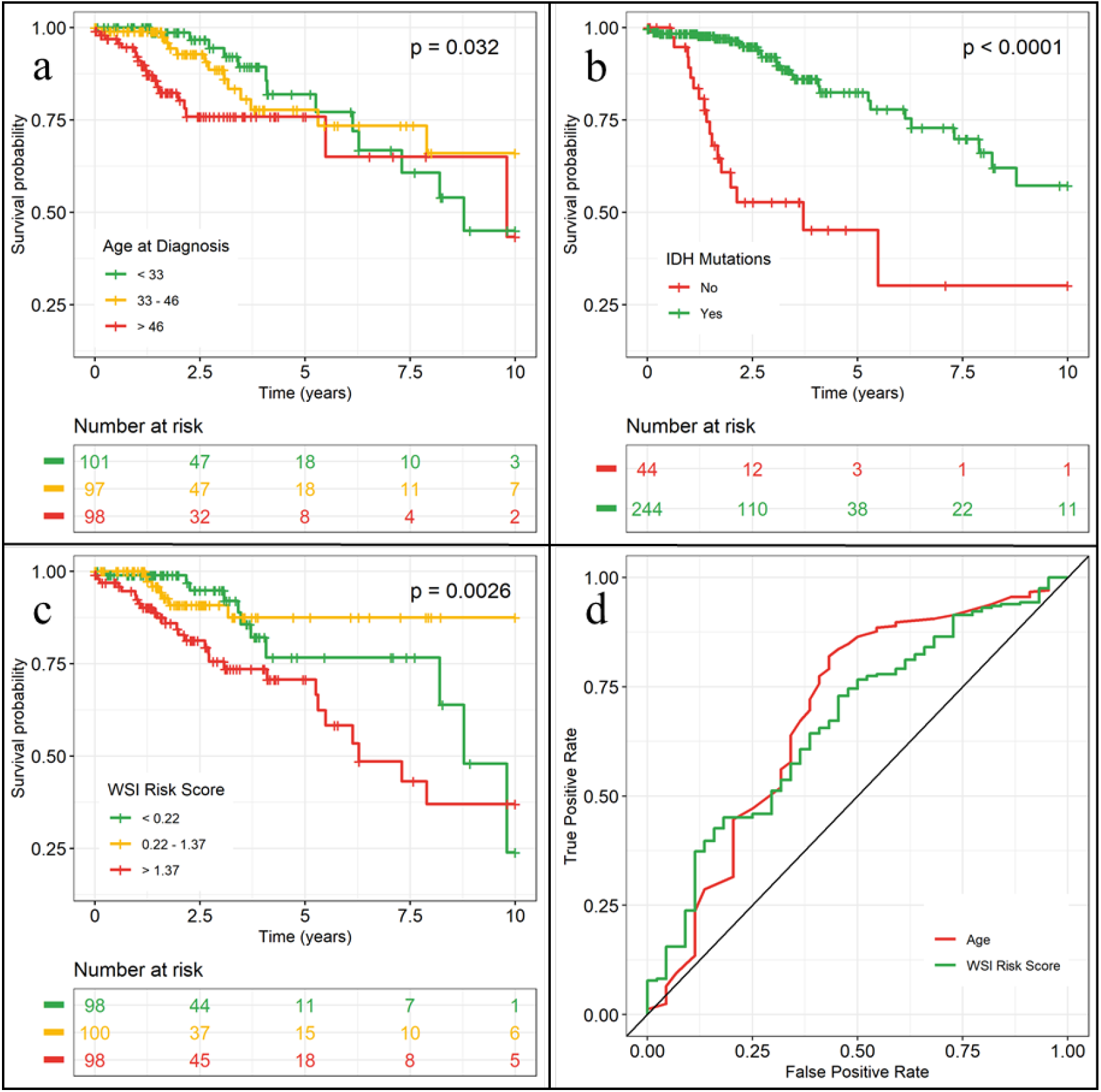
Kaplan-Meier curves and ROC curves evaluated among patients with LGG. **(a), (b), (c)** Kaplan-Meier curves by age, IDH mutations, and WSI risk score, respectively. P-value was calculated by log-rank test. **(d)** ROC curve for IDH mutation prediction.

In the WHO grade 3 cases (Supplementary Table S3), the WSI-based risk scores had lower performance alone (0.654, 95% CI: 0.537, 0.768). Age and IDH mutations together achieved a C-index of 0.786 (95% CI: 0.683, 0.877). When combining WSI risk scores, the model performance improved slightly to 0.792 (0.701, 0.876). Kaplan-Meier curves for patients with WHO grade 3 gliomas are shown in Supplementary Figure S1.

### Model performance and comparison with clinical features for IDH prediction

The AUC of the WSI-based models for prediction of IDH mutations was 0.667 (95% CI: 0.532, 0.784) (Table 1 and Figure 1d). In addition, age is a strong predictor of IDH mutations with AUC of 0.689 (95% CI: 0.552, 0.816). Race is a weak predictor with AUC of 0.567 (95% CI: 0.480, 0.650). Combining race and WSI-based scores, the AUC was increased to 0.687. When combining age and race, the AUC was 0.711 (95% CI: 0.585, 0.834). Including WSI-based scores raised the AUC to 0.739, with 0.028 improvement (95% CI: −0.051, 0.078). For WHO grade 3 cases (Supplementary Table S3 and Supplementary Figure S1), the WSI-based scores can predict IDH mutations with an AUC of 0.814 (95% CI: 0.721, 0.897), which is much higher than the demographic and clinical predictors. When combining age, the AUC was 0.845 (95% CI: 0.759, 0.919), which is a statistically significant improvement over using only age as the predictor (0.122, 95% CI: 0.001, 0.198).

### Prognosis prediction using WSI predicted IDH mutation probability

Additionally, we explored if WSI predicted IDH mutation probability can be used to replace IDH mutation status measurement in predicting prognosis (Table 2). We found predicted IDH mutation probability alone achieved a C-index of 0.727, which is, notably, greater than WSI risk score (0.715) and IDH mutations (0.692). When combining with age, the C-index increased to 0.767 (95% CI: 0.646, 0.862), but not as good as combining age and IDH mutations (C-index = 0.774). Finally, when combining predicted IDH mutation probability with age and WSI risk score, the C-index was 0.771 which was better than age and survival risk score (0.766), but not as good as combining age, survival risk score and IDH mutations (0.784).

**Table 2.**
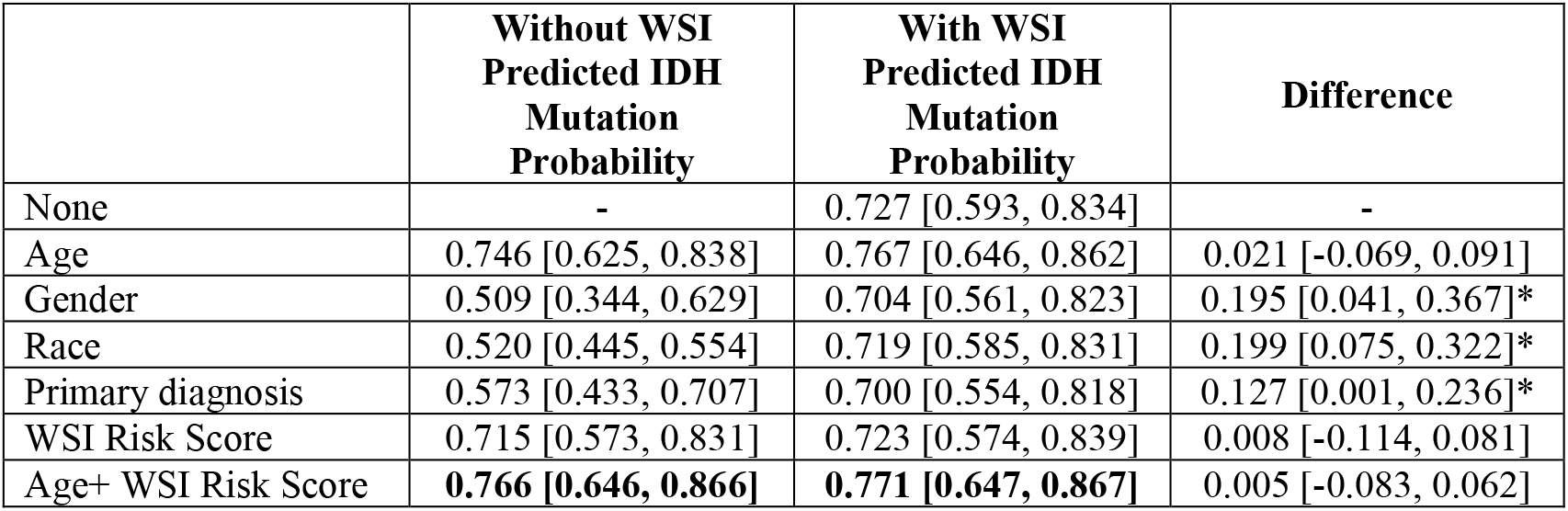
Performance of survival prediction using predicted IDH mutation probability evaluated among patients with LGG. 95% confidence intervals were derived from 10,000 bootstrapping replications. **Bold** texts indicate best performance for each column. * indicates statistically significant difference (p < 0.05).

|  | Without WSI<br>Predicted IDH<br>Mutation<br>Probability | With WSI<br>Predicted IDH<br>Mutation<br>Probability | Difference |
| --- | --- | --- | --- |
| None | - | 0.727 [0.593, 0.834] | - |
| Age | 0.746 [0.625, 0.838] | 0.767 [0.646, 0.862] | 0.021 [-0.069, 0.091] |
| Gender | 0.509 [0.344, 0.629] | 0.704 [0.561, 0.823] | 0.195 [0.041, 0.367]* |
| Race | 0.520 [0.445, 0.554] | 0.719 [0.585, 0.831] | 0.199 [0.075, 0.322]* |
| Primary diagnosis | 0.573 [0.433, 0.707] | 0.700 [0.554, 0.818] | 0.127 [0.001, 0.236]* |
| WSI Risk Score | 0.715 [0.573, 0.831] | 0.723 [0.574, 0.839] | 0.008 [-0.114, 0.081] |
| Age+ WSI Risk Score | **0.766 [0.646, 0.866]** | **0.771 [0.647, 0.867]** | 0.005 [-0.083, 0.062] |

### Visualization of model predictions

The average WSI-based risk scores across patients were 0.947 (standard deviation: 1.587). Prediction results on the whole slide and patch level are shown in Figure 2, and Supplementary Figure S2. Increased tumor cell density and tumor cell atypia, i.e., increased nuclear size, hyperchromasia, and irregular nuclear contours, are associated with higher grade and worse prognosis. The images of the resection specimen in Figure 2a show a diffusely infiltrating glial neoplasm with many areas of high cellularity and pleomorphism. This tumor was diagnosed at the time as an oligoastrocytoma (mixed glioma), and the patient died 1.4 years after diagnosis. The predicted risk score was high (10.77). The histology of this tumor differs dramatically from the one shown in Figure 2b, which reveals only small foci of hypercellularity and atypia (insets, left). Much of the resection specimen from this 32-year-old man diagnosed with mixed glioma showed reactive astrogliosis and mildly infiltrated brain parenchyma (insets, right). The model’s low predicted risk score of 0.65 is consistent with the low grade histologic features of this tumor. The patient’s relatively long survival of six years corroborated the model’s performance.

**Figure 2.**
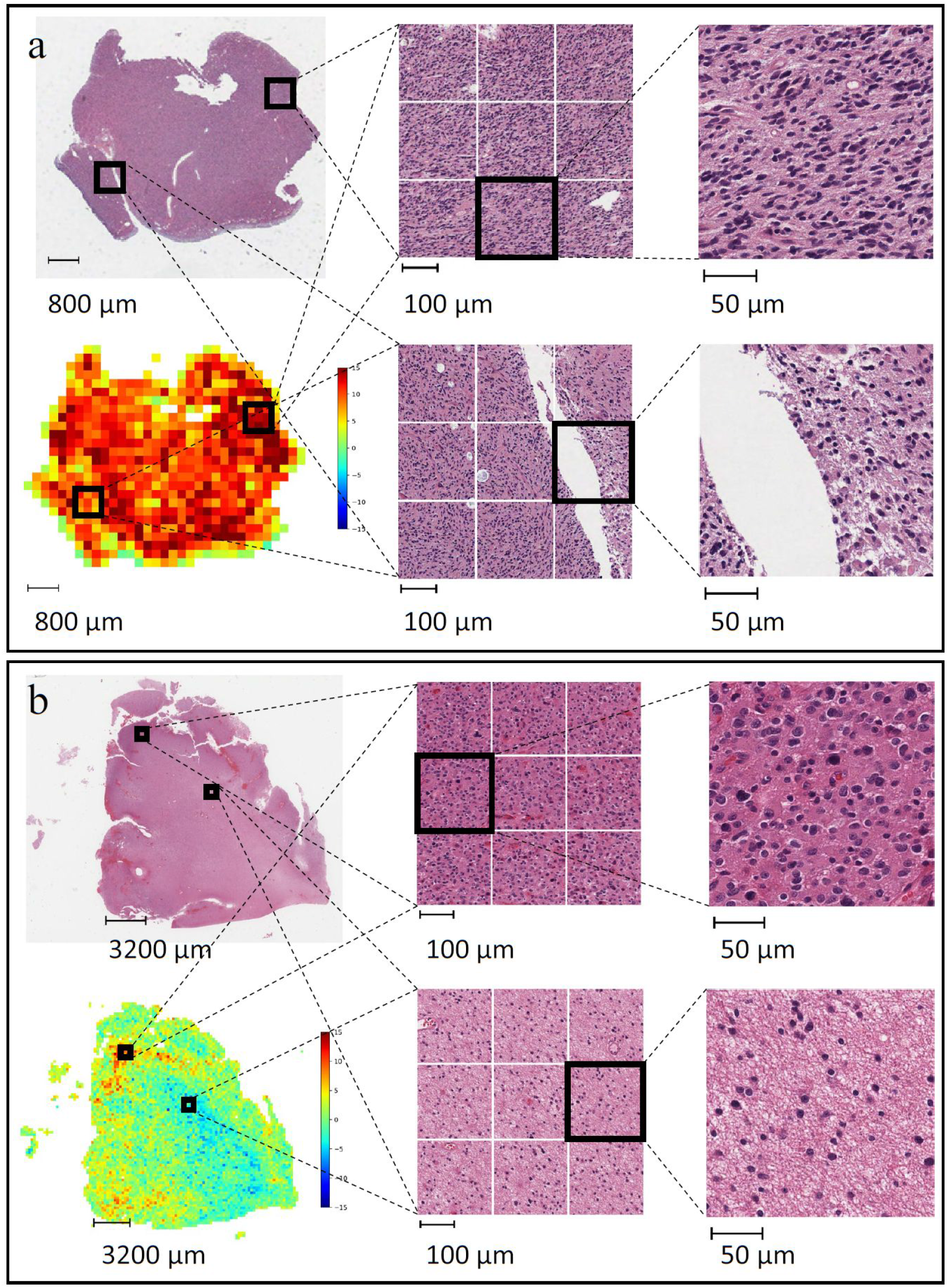
Example predictions on whole slide images for prognosis prediction. **(a)** A 59-year-old female patient diagnosed with mixed glioma, died 1.4 year after diagnosis. The predicted risk score is 10.77. **(b)** A 32-year-old man diagnosed with mixed glioma, died 6 years later. The predicted risk score is 0.65.

## DISCUSSION

In this study, we have shown that by using deep learning models on WSIs, we are able to achieve promising results for predicting prognosis and IDH mutations on the lower-grade gliomas dataset from the TCGA database. The performance of the deep learning model based on WSIs alone is better than the model based on the primary diagnosis and some demographic variables such as race and gender, but not as good as age at diagnosis. Combining WSI-based deep learning predictions with demographic and clinical features could further improve the model performance to up to 0.784 for the prediction of prognosis and 0.739 for the prediction of IDH mutations. We also found if WSI predicted IDH mutation probability is used instead of IDH mutation status measurement, we can still obtain a C-index of 0.771. Our results were further validated using WHO grade 3 patients which were not used during the training and hyperparameter selection.

Previous work on the application of deep learning models to LGG datasets is relatively limited. Studies using less restrictive data inclusion/exclusion criteria reported higher performance in the survival prediction task^11,25^ and IDH mutation prediction task^23^. This lower performance in the stage 2 only patients could be due to the smaller sample size and less variation in the disease severity.

During initial experiments, we noticed that for a single data split, the higher performance in the validation dataset does not necessarily translate to higher performance in the test dataset. This could be due to unbalanced sampling when sample size is small. We also found that a discrepancy in validation and test AUC occurred in Liu et al.’s study^23^. For example, the AUC for their baseline model achieved an AUC of 0.920 on the test set, while in the validation dataset the AUC is 0.823. This highlights the difficulty in obtaining a balanced train/validation/test splitting with limited sample size. Our adoption of the repeated data splits and pooling method can ameliorate this problem.

There are several limitations to this study. First, the sample size in the study is relatively small and the number of lost to follow-up is substantial. With only a total of 296 patients (among which 49 were observed at the endpoint and 44 were IDH wild-type), developing a deep learning framework is challenging. The small sample size also limited the power to detect statistically significant improvement by using the predictions based on WSIs over only demographic and clinical information. Secondly, we did not evaluate the performance of our models on additional datasets; thus, the generalizability of this method needs further validation. Thirdly, the cause of death was not recorded in the TCGA dataset, thus our ground truth might not be accurate for all the samples which could affect the model performance.

Notably, histological information could only explain part of the variance in survival time. Other information, such as the location of the tumor, treatment, and comorbidities are also important determinants of the progression of the disease. In this study, we did not include important clinical data, such as treatment, in our analysis, as the detailed information was not available in the TCGA dataset. In future work, we will pursue expanding our dataset and include this additional relevant information in our analysis. We expect incorporating additional demographic, clinical, and genetic/molecular information in our method could potentially improve the ability to predict prognosis of patients diagnosed with LGG.

## MATERIALS AND METHODS

### Data Source

The digitized hematoxylin and eosin (H&E) stained whole slides used in this project were obtained from The Cancer Genome Atlas (TCGA) database. TCGA database is de-identified and is publicly available on the Web. Therefore, this project does not meet the requirements of human subject research. Only low-grade diffuse gliomas were included in our analysis (number of patients = 307). There are two different types of whole slide images (WSIs) in this dataset, namely formalin-fixed paraffin-embedded (FFPE) slides and frozen section slides^26^. Since the frozen section slides contain many artifacts, we only included FFPE slides in our dataset (number of patients = 296, number of WSIs = 524).

Demographics and clinical information were also downloaded from the TCGA website. For the deceased patient, the follow-up time was derived from “days to death”. For patients who were alive at the last follow-up, the follow-up time was derived from “days to last follow-up”. IDH mutation status was derived from IDH1 and IDH2 mutations variables. Eight participants without IDH mutation information were excluded from IDH related analysis. Demographic and clinical information including age, gender, race, and primary diagnosis, was used in our analysis for comparison purposes. The average age of the patients in our dataset was 40.9 years with a standard deviation of 13.0 years. Among those, 55.7 percent were men, and the majority (91.6%) of the patients were white. The proportions of patients diagnosed as astrocytoma, oligoastrocytoma, and Oligodendroglioma were 19.9%, 43.9% and 36.1%, respectively. 80.2% of the patients had an IDH1 mutation, while 4.5% of the participants had an IDH2 mutation (Supplementary Table S1).

To further evaluate the performance of our method, we obtained the grade 3 glioma cases from the TCGA database for testing purposes only (number of patients = 194, number of whole slides images = 319). The data processing procedure for grade 3 cases is the same as the grade 2 cases. The distribution of demographic variables was similar to grade 2 cases, and IDH mutations were present in 68.6% of the cases (Supplementary Table S2).

### Preprocessing of whole slide images

As WSIs are large in size and cannot fit in GPU memory, several preprocessing steps were taken to extract patches from the original images. We loaded the WSIs at the magnification factor of 10× (1μm/pixel) and extracted patches with a size of 224×224 pixels without overlap. Background patches were excluded by using color thresholding. A total of 1,887,767 patches were generated through this process.

### Model Architecture

For the prognosis prediction task, our model architecture is adapted from the proposed work by Wulczyn et al.^24^ and is illustrated in Figure 3. In summary, for each batch, *n* participants were randomly chosen from the training dataset. For each participant, *k* patches were randomly selected. These patches were fed into a deep learning model. The ResNet-18 model with pre-trained ImageNet weights was used as the backbone model^27^, and a fully-connected layer was replaced by an identity layer. The output size for each patch was 512. We then averaged the feature vectors over *k* patches for each participant and used the pooled features for risk estimation through a subsequent two-layer neural network with 128 neurons and 1 neuron for each layer, respectively. The final output can be interpreted as risk scores and the loss is calculated as the negative log Cox partial likelihood, which is defined as

**Figure 3.**
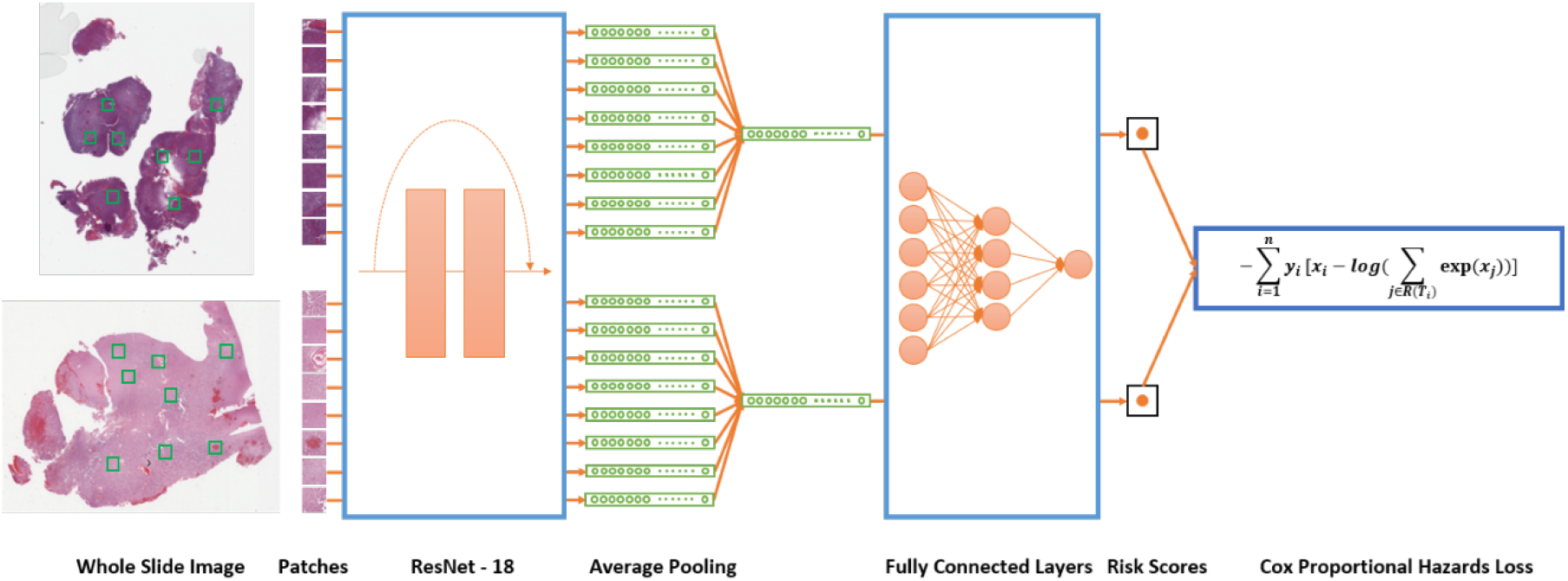
An overview of the deep learning pipeline for prognosis prediction. Patches of size 224×224×3 are randomly sampled from whole slide images at a 10× magnification level. The ResNet-18 Convolutional Neural Network transformed each patch into a 512×1 vector. Average pooling is performed at the patient level. The patient level vectors then go through a two-layer fully connected network with final output size of 1, which can be interpreted as risk scores. Cox proportional hazards loss is calculated using the risk scores with consideration of follow-up time and vital status. The gradient is calculated and backpropagated through the fully connected layers and the ResNet-18 layers to train the entire model.

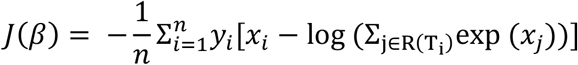

where *n* is the number of patients, *x*_*i*_ is the risk score, *y*_*i*_ is the event indicator (0 for alive and 1 for death), *R*(*T*_*i*_) is the risk set at the event time of *i*th patient.

The model architecture for the binary IDH mutation prediction task is similar to the one for prognosis prediction, except that the final output size is 2. Since the percentage of participants with IDH mutations was much larger than that of participants without IDH mutations, we used weighted cross-entropy loss to handle the imbalanced dataset by assigning a larger weight to cases without IDH mutations.

During validation, 100 random patches were selected for each patient in the validation group for a balance between variations and efficiency. All the patches were used when making out-of-sample predictions for cases in the test set.

### Model evaluation metrics

Concordance index (C-index), which is defined as the proportion of concordant pairs among all possible pairs, was used as the evaluation metric of our prognosis prediction model. Area under ROC (receiver operating characteristic) curve (AUC) was used as the evaluation metric for the binary classification tasks.

### Training-validation data splits

The data splitting was performed at the patient level to avoid the information leak across partitions. Due to limited training data, to ensure more balanced group splits, we first sorted the patients by vital status and follow-up time, then created multiple 4-patient-blocks. Within each block, we assigned 2 patients to the training group and 1 patient to each of the validation and test groups. This random splitting was repeated 8 times for the purpose of hyperparameter tuning, and was repeated another 24 times for model evaluation (as explained below).

### Hyperparameter tuning

Within each random data split, our deep learning model was fit on the training split, with its performance monitored using the validation split. When the training is finished, out-of-sample prediction was obtained for the test dataset. We repeated this process in all of these 8 repetitions and used the average validation performance metrics to choose the best set of hyperparameters. The final set of hyperparameters chosen was batch-size of *n* = 8 patients with *k* = 8 patches for each individual (64 patches per batch in total), initial learning rate of 1e-4 for the fully connected layers and 3e-7 for the convolutional layers.

Data augmentation methods, such as random horizontal and vertical flips and color jittering, were used during training time. To mitigate overfitting, we applied an L1 penalty with regularization strength of 0.01 on the fully connected layers. Adam optimization was used for training. Cosine annealing was used as the learning rate scheduler. Each model was evaluated when every 20,480 (i.e., 320 steps) patches were used and training was stopped after 96 thousand steps.

### Bootstrapping on out-of-sample predictions

After training and hyperparameter tuning across the first 8 random splits, we trained the models with the same hyperparameters using the additional 24 random splits. These 32 models provided 32 out-of-sample predictions. The test set size for each model was one fourth of the total dataset. Because each participant was selected into the test dataset with a probability of 0.25, the number of out-of-sample predictions for a participant follows the Poisson distribution with a mean of 8 (min = 2, max = 18). We ensembled all the out-of-sample predictions by averaging them as the final prediction.

Subsequently, we performed a bootstrapping method to evaluate the model performance. To do so, we randomly selected 296 observations from the entire dataset with replacement as the training dataset (about 63% of the patients). A statistical model (Cox or logistic) using demographic and clinical information with/without deep learning predictions was fit on the training dataset. The participants who were not selected formed the test dataset (about 37% of the patients) and were used to evaluate the performance of the statistical model. We repeated this process 10,000 times to estimate the distribution of C-index and AUC without or with deep learning predictors as well as their difference. The deep learning framework was implemented in PyTorch (version 1.1.0). The statistical tests were performed using R (version 3.6.1).

### Results visualization

Kaplan-Meier curves were used to present the observed survival probability over time by tertiles of age and WSI risk score, and IDH mutation status. The Kaplan-Meier curves were replicated for patients with grade 3 gliomas using the same cut-offs. For the IDH mutation prediction task, ROC curves were plotted with age and WSI-based IDH mutation probability as the predictor.

To visualize the model performance at the whole slide level, we selected two patients from prognosis prediction task and another two from IDH mutation prediction task. For the prognosis prediction task, we chose one patient who died shortly after diagnosis, and another patient who survived at least 5 years after diagnosis. For the IDH mutation prediction task, we chose one patient with an IDH mutation and one without. One whole slide image was selected for each patient. Representative regions from the slide were selected for a more detailed view.

## Data Availability

This project's source of data is the TCGA database, which is publicly available on the Web.

https://portal.gdc.cancer.gov/projects/TCGA-LGG

## Data availability

This project’s source of data is the TCGA database, which is publicly available on the Web (https://portal.gdc.cancer.gov/projects/TCGA-LGG).

## AUTHOR CONTRIBUTIONS

Concept and design: G.J.Z. and S.H.; Acquisition, analysis, or interpretation of data: S.J.; Drafting of the manuscript: S.J.; Critical revision of the manuscript for important intellectual content: All authors.; Statistical analysis: S.J.; Obtained funding: S.H.; Administrative, technical, or material support: S.H.; Supervision: S.H.

