## Supplementary materials for "Predicting Prognosis and IDH Mutation Status for Patients with Lower-Grade Gliomas Using Whole Slide Images"

**Figure S1. Kaplan-Meier curves and ROC curve evaluated among patients with grade 3 gliomas. (a), (b), (c)** Kaplan-Meier curves by age, IDH mutations, and WSI risk score, respectively. P-value was calculated by log-rank test. **(d)** ROC curves for IDH mutation prediction.

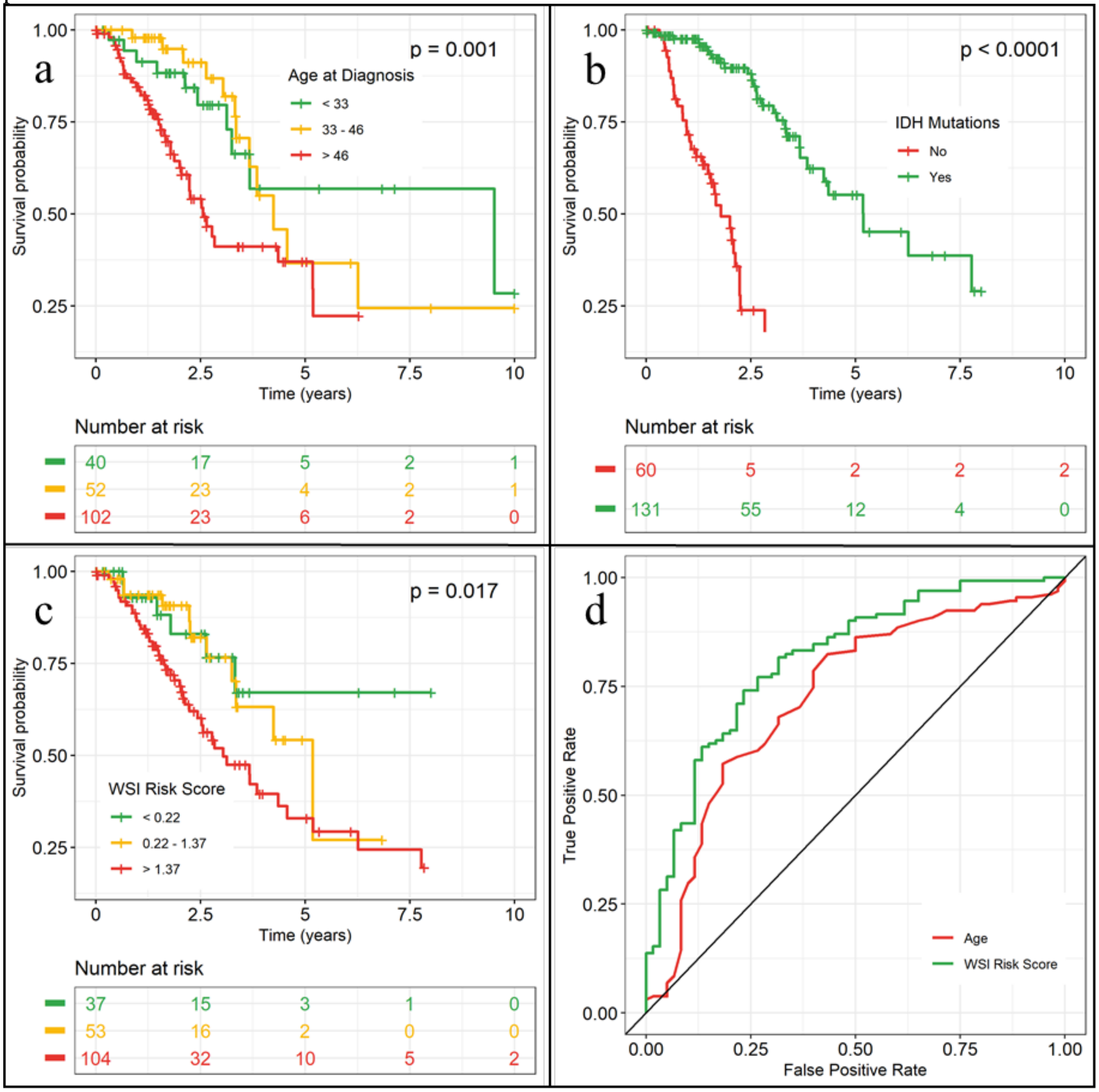

**Figure S2. Example predictions on whole slide images for IDH mutation prediction. (a)** A 33-year-old woman diagnosed with mixed glioma with an IDH mutation. **(b)** A 61-year-old woman diagnosed with mixed glioma without an IDH mutation.

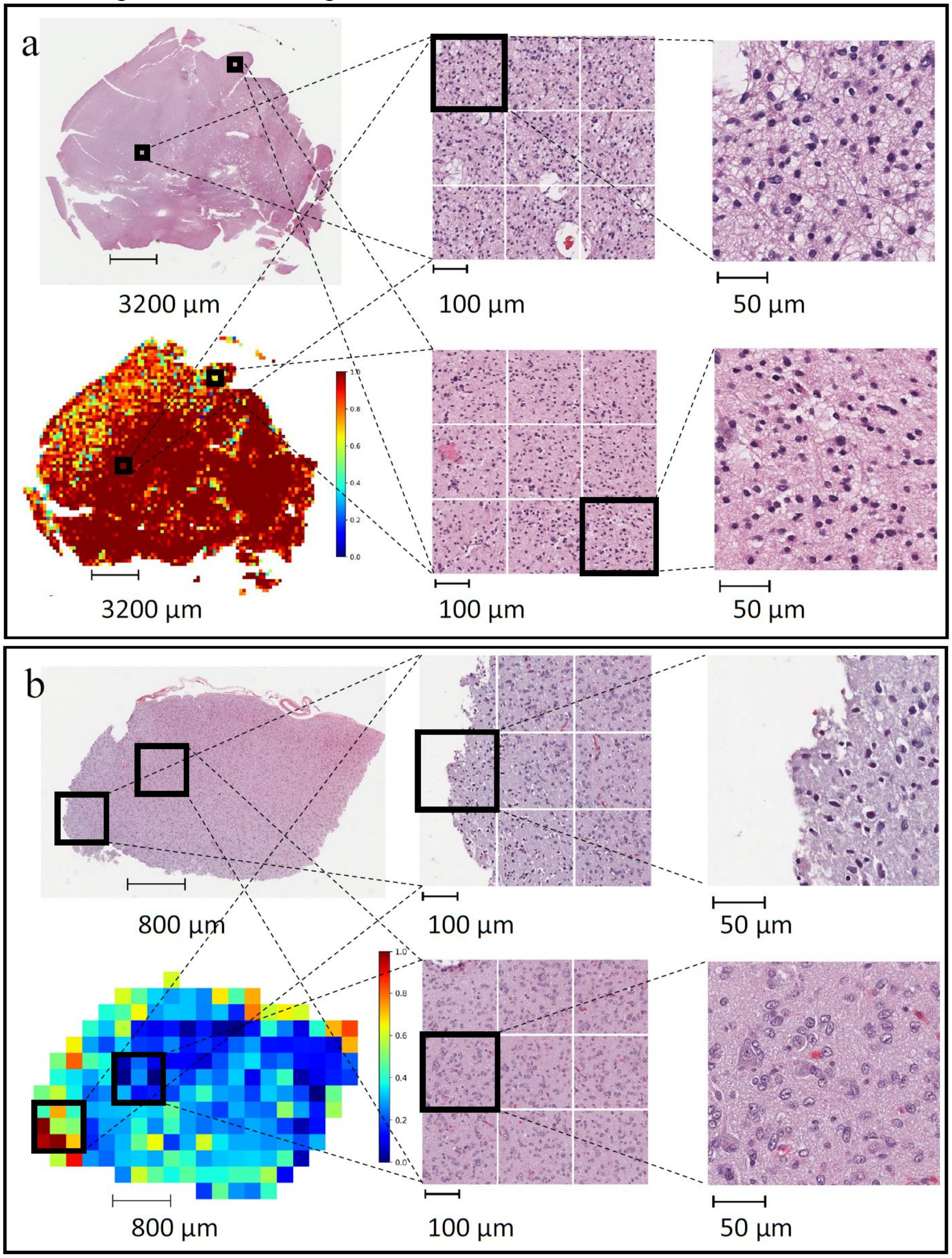

**Table S1. Distribution of baseline characteristics among patients with LGG.**

| **Variables** | **Mean (SD) / N (%)** |
|---|---|
| Age | 40.9 (13.0) |
| Gender | |
| Male | 165 (55.7%) |
| Female | 131 (44.3%) |
| Race | |
| White | 271 (91.6) |
| Black or African American | 13 (4.4%) |
| Asian | 5 (1.7%) |
| American Indian or Alaska native | 1 (0.3%) |
| Not reported | 6 (2.0%) |
| Primary diagnosis | |
| Astrocytoma | 59 (19.9%) |
| Oligoastrocytoma (Mixed Glioma) | 130 (43.9%) |
| Oligodendroglioma | 107 (36.1%) |
| IDH mutations | |
| IDH1 mutation | 231 (80.2%) |
| IDH2 mutation | 13 (4.5%) |
| Either IDH1 or IDH2 mutation | 244 (84.7%) |

**Table S2. Distribution of baseline characteristics among patients with grade 3 gliomas.**

| **Variables** | **Mean (SD) / N (%)** |
|---|---|
| Age | 46.6 (13.3) |
| Gender | |
| Male | 106 (54.6%) |
| Female | 88 (45.4%) |
| Race | |
| White | 181 (93.3%) |
| Black or African American | 7 (3.6%) |
| Asian | 2 (1.0%) |
| American Indian or Alaska native | - |
| Not reported | 4 (2.1%) |
| Primary diagnosis | |
| Anaplastic Astrocytoma | 122 (62.9%) |
| Anaplastic Oligodendroglioma | 72 (37.1%) |
| IDH mutations | |
| IDH1 mutation | 125 (65.4%) |
| IDH2 mutation | 6 (3.1%) |
| Either IDH1 or IDH2 mutation | 131 (68.6%) |

**Table S3. Model performance statistics for survival prediction task and IDH mutation prediction task, evaluated among patients with grade 3 gliomas.** 95% confidence intervals were derived from 10,000 bootstrapping replications. **Bold** texts indicate best performance for each column. * indicates statistically significant difference ($p < 0.05$).

| **Survival prediction performance: C-index [95% CI]** | | | |
|---|---|---|---|
| | **Without WSI Risk Score** | **With WSI Risk Score** | **Difference** |
| None | - | 0.654 [0.537, 0.768] | - |
| Age | 0.729 [0.616, 0.833] | 0.758 [0.660, 0.845] | 0.029 [-0.041, 0.082] |
| Gender | 0.514 [0.370, 0.613] | 0.652 [0.536, 0.757] | 0.138 [-0.006, 0.287] |
| Race | 0.501 [0.445, 0.550] | 0.644 [0.528, 0.760] | 0.143 [0.028, 0.259]* |
| Primary diagnosis | 0.563 [0.375, 0.646] | 0.661 [0.544, 0.775] | 0.098 [-0.014, 0.286] |
| IDH mutations | 0.724 [0.640, 0.803] | 0.757 [0.657, 0.844] | 0.033 [-0.040, 0.092] |
| Age+IDH mutations | **0.786 [0.683, 0.877]** | **0.792 [0.701, 0.876]** | 0.006 [-0.035, 0.037] |
| **IDH mutation prediction performance: AUC [95% CI]** | | | |
| | **Without WSI Predicted IDH Mutation Probability** | **With WSI Predicted IDH Mutation Probability** | **Difference** |
| None | - | 0.814 [0.721, 0.897] | - |
| Age | **0.728 [0.619, 0.833]** | **0.845 [0.759, 0.919]** | 0.117 [0.001, 0.198]* |
| Gender | 0.521 [0.417, 0.618] | 0.807 [0.713, 0.893] | 0.286 [0.146, 0.415]* |
| Race | 0.499 [0.440, 0.545] | 0.812 [0.708, 0.901] | 0.313 [0.210, 0.415]* |
| Primary diagnosis | 0.646 [0.556, 0.732] | 0.834 [0.740, 0.915] | 0.188 [0.091, 0.264]* |
| Age + Race | 0.716 [0.600, 0.821] | 0.842 [0.739, 0.917] | 0.126 [0.014, 0.211]* |
